# Rasch Validation of the Revised Body Awareness Rating Questionnaire (BARQ-R) in Adults with Musculoskeletal Pain, Adults with Spinal Cord Injury, and Community-Dwelling Adults in the US

**DOI:** 10.1101/2022.04.19.22274054

**Authors:** Sydney Carpentier, Wei Deng, Jena Blackwood, Ann Van de Winckel

## Abstract

**Background:** To establish Rasch validation of the Revised Body Awareness Rating Questionnaire (BARQ-R) in adults with musculoskeletal pain, community-dwelling adults without pain, and adults with spinal cord injury (SCI) who have neuropathic pain.

**Materials and Method:** The BARQ-R has 12 items with scores ranging from 0 (completely disagree) to 3 (completely agree). Through Rasch analysis, we evaluated unidimensionality through item and person fit, targeting of the population, person separation reliability (PSR), local item dependence (LID), and principal components analysis of residuals (PCAR).

**Results:** The BARQ-R in adults with musculoskeletal pain (n=152; average age = 52.26±16.13 years) showed good targeting (person mean location: -0.36±0.88 logits), minimal floor effect (0.01%), and no ceiling effect (0.00%) and had good reliability (PSR=0.75). The BARQ-R in community-dwelling adults (n=471; average age = 49.63±17.57 years) had a person mean location of -0.62±1.09 logits, minimal floor (2.63%), and minimal ceiling effect (0.43%) after rescoring 2 items and deleting 3 items and had good reliability (PSR=0.74). The BARQ-R in adults with SCI-related neuropathic pain (n=44; average age = 55.45±13.47 years) showed good targeting after rescoring 7 items (person mean location: -0.33±0.71 logits), no floor effect (0.00%) or ceiling effect (0.00%) but had poor reliability (PSR=0.65).

**Conclusions:** The BARQ-R shows sufficient fit to be used in clinical settings for group decision-making for both adults with musculoskeletal pain and community-dwelling adults. However, in adults with SCI-related neuropathic pain, preliminary Rasch analysis of the BARQ-R showed low reliability and therefore the BARQ-R is not recommended for clinical use in that population. Validation in larger groups of adults with SCI as well as more diverse samples are needed.

## INTRODUCTION

Body awareness is generally defined as the ability to understand and become aware of (i) how the body is situated in space (i.e., proprioception); sensory signals, such as visual, auditory, tactile signals that are perceived in that space (i.e., exteroception); and internal bodily states (i.e., interception).^1–9^ This definition encompasses awareness of body posture and body movements, which are elements that are addressed in physical therapy.^10^ Mind and body approaches such as Tai Chi, Qigong, Yoga as well as other meditative practices such as mindfulness, breathing exercises, and even physical activity impact body awareness,^11,12^ necessitating the use of a scale that can measure a person’s body awareness ability.

To measure body awareness, Dragesund *et al*. (2010) developed a patient-reported outcome measure, the Body Awareness Rating Questionnaire (BARQ), intended for adults with chronic musculoskeletal pain and psychosomatic disorders.^13^ In the context of this scale, body awareness was defined as a person’s ability to sense muscle tension and body movements as well as reflect on their emotions and attitudes toward their own body.^11^ People who are aware of their body posture and muscle tension will readjust their posture more quickly,^13^ thereby avoiding muscle tension and resultant pain. Items of the BARQ were developed through interviewing therapists specializing in Norwegian Psychomotor Physiotherapy (NPMP) ─a mind-body physiotherapeutic approach for improving pain levels and function ^13,14^─ and through focus groups of individuals with chronic pain who both received and were on a waiting list to receive the NPMP therapy. This resulted in 66 items. Through Cronbach’s *α* test and exploratory factor analysis, the resulting scale consisted of 24 items with 4 subscales reflecting separate aspects of body awareness: Function, Mood, Feelings, and Awareness. The BARQ is scored using a 7-point Likert scale ranging from 1 (totally agree) to 7 (totally disagree).

To assess whether the BARQ has satisfactory measurement properties, Dragesund *et al*. (2012) tested its validity and reliability in a sample of Norwegian adults with long-lasting musculoskeletal pain and/or psychosomatic disorders. Reliability was satisfactory for all subscales; however, construct and discriminate validity was only substantial for three subscales (i.e., Function, Feelings, and Awareness), but was not substantial for the Mood subscale.

As a next step, Dragesund *et al*. (2018) evaluated the structural validity of the BARQ through Rasch analysis in 48 adults with musculoskeletal pain, which resulted in the Revised BARQ (BARQ-R)^15^ with 12 items reflecting statements on body awareness, on a 4-point ordinal scale ranging from 0 (completely disagree) to 3 (completely agree), with higher scores reflecting lower degrees of body awareness.^15^

It is critical to replicate these findings in both a larger sample and in other populations outside of Norway. In addition to pain populations, it is important to determine what the typical range of BARQ-R scores is in adults in a community who do not have pain. Also, it is important to know if the BARQ-R scale can be used in other patient populations known to have body awareness deficits,^16,17^ such as adults with spinal cord injury (SCI) and neuropathic pain.

Therefore, the objectives of this study were to (1) validate the results from Dragesund *et al*. (2018) by evaluating the structural validity of the BARQ-R with Rasch analysis in a large sample of adults with self-reported musculoskeletal pain in the United States (US), (2) evaluate the scoring range of the BARQ-R in community-dwelling adults in the US who report no pain, and (3) assess the BARQ-R as a pilot Rasch analysis in adults with SCI who had neuropathic pain.

## MATERIALS AND METHODS

### Participants

Participants were recruited at the Minnesota State Fair, Highland Fest, from a contact list of the Brain Body Mind Lab at the University of Minnesota, and through flyers and website announcements. Participants must be 18-75 years of age to participate in the study. Those who were pregnant, who did not speak English, and/or those who could not provide consent were excluded from the study.

This study adhered to the principles of the most recent Declaration of Helsinki (2013) and received approval from the Institutional Review Board of the University of Minnesota (IRB#s 00008476; 00011997; 00005656; 00005849). All participants provided informed consent. Adults with SCI-related neuropathic pain completed the BARQ-R as part of a clinical trial. They signed consent on the secure University of Minnesota REDCap platform. All other participants completed the BARQ-R as part of an anonymous questionnaire and, thus, we did not collect any personal or identifiable information (including signatures) from those participants. After they read and provided consent, participants were quizzed on the comprehension of the content of the consent form through the University of California, San Diego Brief Assessment of Capacity to Consent (UBACC)^18^. All information was stored automatically on the secure University of Minnesota REDCap platform.

### Main outcome measures

The BARQ-R is a patient-reported outcome measure consisting of 12 items, scored on a 4-point ordinal scale, ranging from 0 (completely disagree) to 3 (completely agree), with higher scores reflecting lower degrees of body awareness.

### Statistical analysis: Rasch Measurement Theory

The structural validity of the BARQ-R was assessed with the Rasch Measurement Theory using the Rasch Unidimensional Measurement Model (RUMM2030) software and was reported following the Rasch Reporting Guideline for Rehabilitation Research (RULER).^19,20^ The Rasch model is a probabilistic model, which states that a person with a higher ability on a certain construct would have a higher probability of obtaining a better score on the scale measuring that construct.^21^

To assess the structural validity of the BARQ-R, we evaluated: (1) the presence of reversed thresholds; (2) item and person fit, with good item fit reflected by fit residuals between -2.50 and +2.50;^21^ (3) targeting of the population, with good targeting reflected by the average person location being within 0.5 logits of average item location and less than 15% floor and ceiling effect,^22^ and (4) personal separation reliability (PSR), which reflects how well we can distinguish groups of persons with different levels of ability,^23^ with ≥ .70 sufficient for group decisions and ≥ .90 sufficient for individual decisions.^19,20^ The interpretation of this value is sometimes compared to Cronbach’s alpha but the error variance structure of PSR and Cronbach’s alpha are reversed.^23,24^

We also evaluated: (5) Local item dependence (LID) measured through residual correlations, with those at least *r* = 0.20 above the average correlation indicating pairs of items that are more correlated with each other, and thus sharing more content than with other items in the scale; and (6) unidimensionality tested through Principal Component Analysis of Residuals (PCAR)^22^ showing unidimensionality if the eigenvalue is < 2.00, and the percent variance on the first residual factor is <10%. Furthermore, a paired sample *t*-test of two subsets of items (correlation with the first principal component loadings of *r* >0.3 or <0.3) showing <5% of significance confirms unidimensionality. The analysis results in a hierarchical order of items ranging from easiest to hardest item. For analyses revealing good structural validity, we provide a score-to-measure table relating the ordinal total score to the Rasch-based logit score and transformed logit score to 0-100% for easier use in the clinic or research. Of note, these score-to-measure tables can only be used for full datasets.

## RESULTS

### Demographic data

There were 667 participants in total: 152 adults with musculoskeletal pain (average age=52.26±16.13 years), 44 adults with SCI-related neuropathic pain (average age=55.45±13.47 years), and 471 community-dwelling adults with no pain (average age=49.63±17.57 years). More details on the demographic data are displayed in **Table 1**.

**Table 1.** Demographics of the study sample.

|  | **Community-**<br>**dwelling**<br>**(n=471)** | **Musculo-**<br>**skeletal pain**<br>**(n=152)** | **SCI and**<br>**neuropathic**<br>**pain (n=44)** | **Total**<br>**(n=667)** |
| --- | --- | --- | --- | --- |
| **Age (years, Mean±SD)** | 49.63±17.57 | 52.26±16.13 | 55.45±13.47 | 50.61±17.1 |
| **Sex (n)** |  |  |  |  |
| Male | 33.97 | 23.03 | 70 | 36.28 |
| Female | 66.03 | 76.97 | 30 | 63.72 |
| Other | 0.21 | 0.00 | 0.00 | 0.15 |
| **Racial background (%)** |  |  |  |  |
| Asian | 2.12 | 5.26 | 2.27 | 5.55 |
| Black/ African American | 1.91 | 2.63 | 2.27 | 2.10 |
| White | 88.54 | 88.16 | 93.18 | 89.00 |
| Multi-racial | 2.12 | 0.66 | 0.00 | 1.65 |
| Hawaiian/other Pacific Islander | 0.00 | 0.00 | 0.00 | 0.00 |
| American Indian/Alaskan | 2.27 | 1.32 | 0.00 | 0.00 |

### Rasch Measurement Theory

All iteration steps are shown in **Table 2**.

**Table 2.**
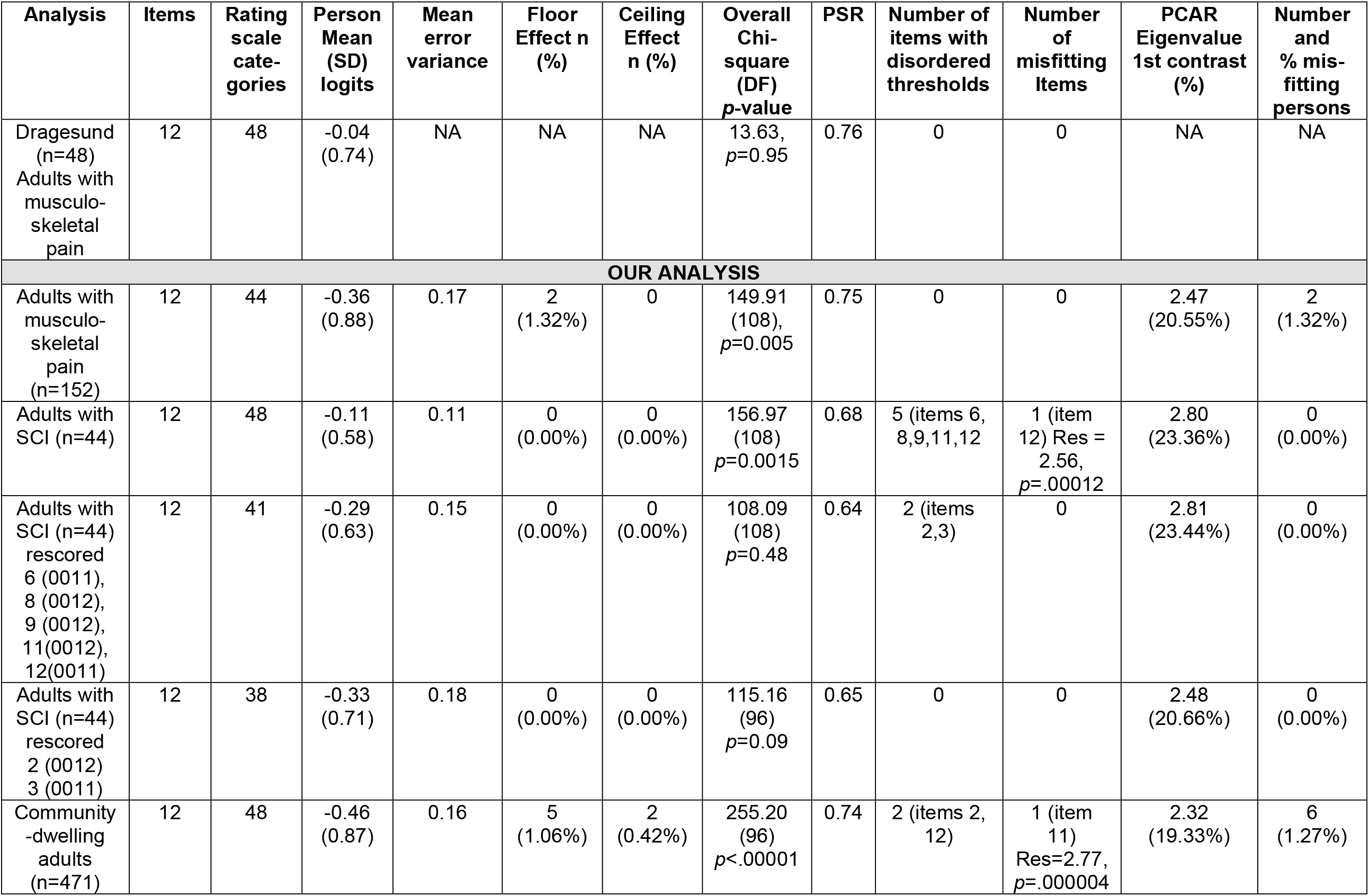
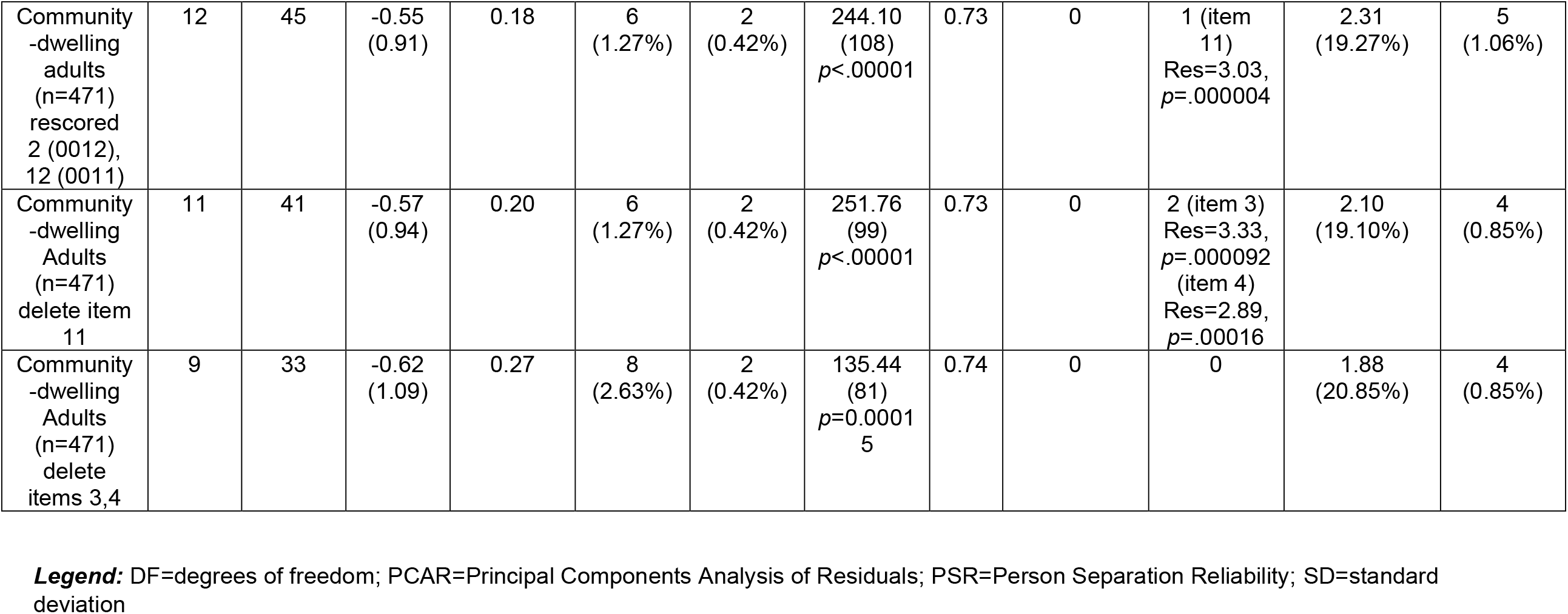
Iteration table.

### Adults with musculoskeletal pain

No items showed disordered thresholds. There were no misfitting items and only 2 (1.32%) misfitting persons. The BARQ-R showed good targeting for adults with musculoskeletal pain (person mean location: -0.36±0.88 logits, **Fig. 1A**), minimal floor effect (0.01%), and no ceiling effect (0.00%). Reliability was good (PSR=0.75). LID was found in 6 item pairs (**Table 3**). PCAR had an eigenvalue of 2.47 (percent variance 20.55%) but the paired *t*-test resulted in only 13.82% of persons having a significantly different logit location on the two subtests, created by the positive and negative principal component loadings. This result is indicative of the scale being multidimensional. “*My body is affected by how I feel*” was the easiest item and “*I don’t like to be touched*” was the hardest item for this group (**Fig. 2A**). The score-to-measure table is provided in **Table 4**.

**Table 3:** Residual Correlation by group.

| Items | Correlation |
| --- | --- |
| **Musculoskeletal Pain $r = 0.29$ (correlation shown is +0.2 above average)** |  |
| 3. I am not aware of the way I breathe.<br>1. I am often tense. | -0.34 |
| 6. My body is tense without me knowing why.<br>1. I am often tense. | 0.31 |
| 11. I avoid paying too much attention to my body.<br>1. I am often tense. | -0.33 |
| 3. I am not aware of the way I breathe.<br>2. My body is affected by how I feel. | -0.30 |
| 3. I am not aware of the way I breathe.<br>4. I don't pay attention to the way I move. | 0.55 |
| 11. I avoid paying too much attention to my body.<br>5. I struggle to relax. | -0.30 |
| **SCI-related neuropathic pain $r = 0.28$ (correlation shown is +0.2 above average)** |  |
| 4. I don't pay attention to the way I move<br>2. My body is affected by how I feel | -0.38 |
| 8. My digestion is affected by how I feel<br>2. My body is affected by how I feel | 0.36 |
| 3. I am not aware of the way I breathe<br>4. I don't pay attention to the way I move | 0.29 |
| 5. I struggle to relax<br>3. I am not aware of the way I breathe | -0.33 |
| 9. I can't get comfortable when I'm lying down<br>3. I am not aware of the way I breathe | -0.29 |
| 10. My body is unpredictable<br>3. I am not aware of the way I breathe | -0.33 |
| 4. I don't pay attention to the way I move<br>5. I struggle to relax | -0.42 |
| 10. My body is unpredictable<br>4. I don't pay attention to the way I move | -0.57 |
| 11. I avoid paying too much attention to my body<br>5. I struggle to relax | -0.37 |
| 12. I don't like to be touched<br>10. My body is unpredictable | -0.28 |

**Table 4.** Rasch-based score-to-measure tables for adults with musculoskeletal pain.

| Total<br>BARQ-R<br>Score | Logit | Converted<br>logits to 0-100 |
| --- | --- | --- |
| 0 | -3.95 | 0.00 |
| 1 | -3.15 | 15.94 |
| 2 | -2.60 | 26.79 |
| 3 | -2.24 | 34.16 |
| 4 | -1.95 | 39.85 |
| 5 | -1.71 | 44.59 |
| 6 | -1.51 | 48.66 |
| 7 | -1.33 | 52.32 |
| 8 | -1.16 | 55.65 |
| 9 | -1.00 | 58.75 |
| 10 | -0.86 | 61.69 |
| 11 | -0.72 | 64.48 |
| 12 | -0.58 | 67.20 |
| 13 | -0.45 | 69.84 |
| 14 | -0.32 | 72.43 |
| 15 | -0.19 | 74.99 |
| 16 | -0.06 | 77.55 |
| 17 | 0.07 | 80.10 |
| 18 | 0.20 | 82.70 |
| 19 | 0.33 | 85.32 |
| 20 | 0.46 | 88.01 |
| 21 | 0.60 | 90.81 |
| 22 | 0.75 | 93.71 |
| 23 | 0.90 | 96.76 |
| 24 | 1.06 | 100.00 |

**Fig. 1.**
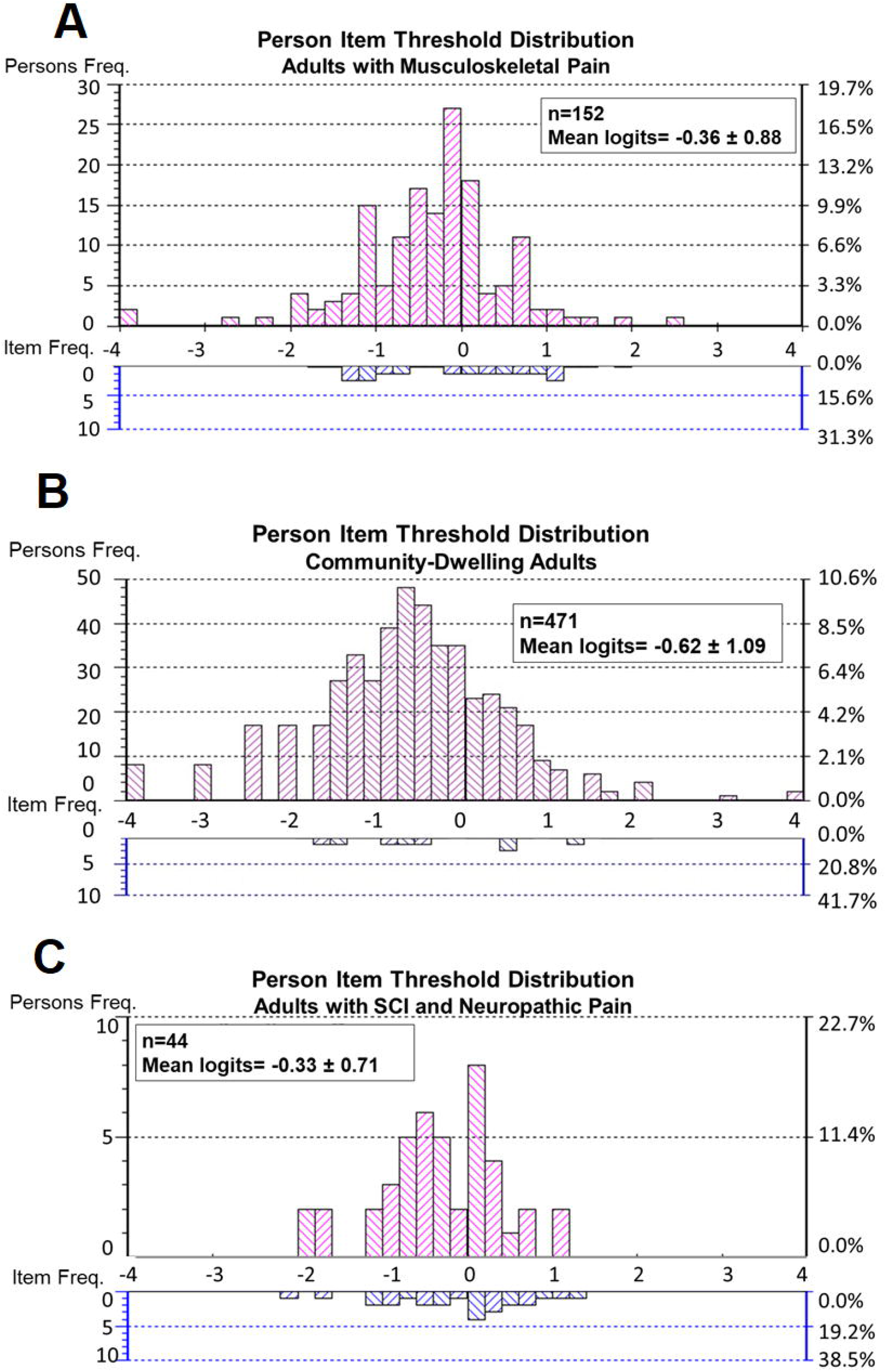
Person-item threshold distribution maps. ***Legend:*** The histogram shows **A:** the frequency of adults with musculoskeletal pain at their level of body awareness ability, **B:** the frequency of community-dwelling adults at their level of body awareness ability, **C:** the frequency of adults with SCI and neuropathic pain at their body awareness ability. Body awareness ability ranges from a high (lowest logit value on the left side of the scale) to low (highest logit value on the right side of the scale). At the bottom of the figure, the blue histogram represents the frequency of item threshold. Following the same logit scale, the easiest items are shown on the left and the hardest items are shown on the right.

**Fig. 2.**
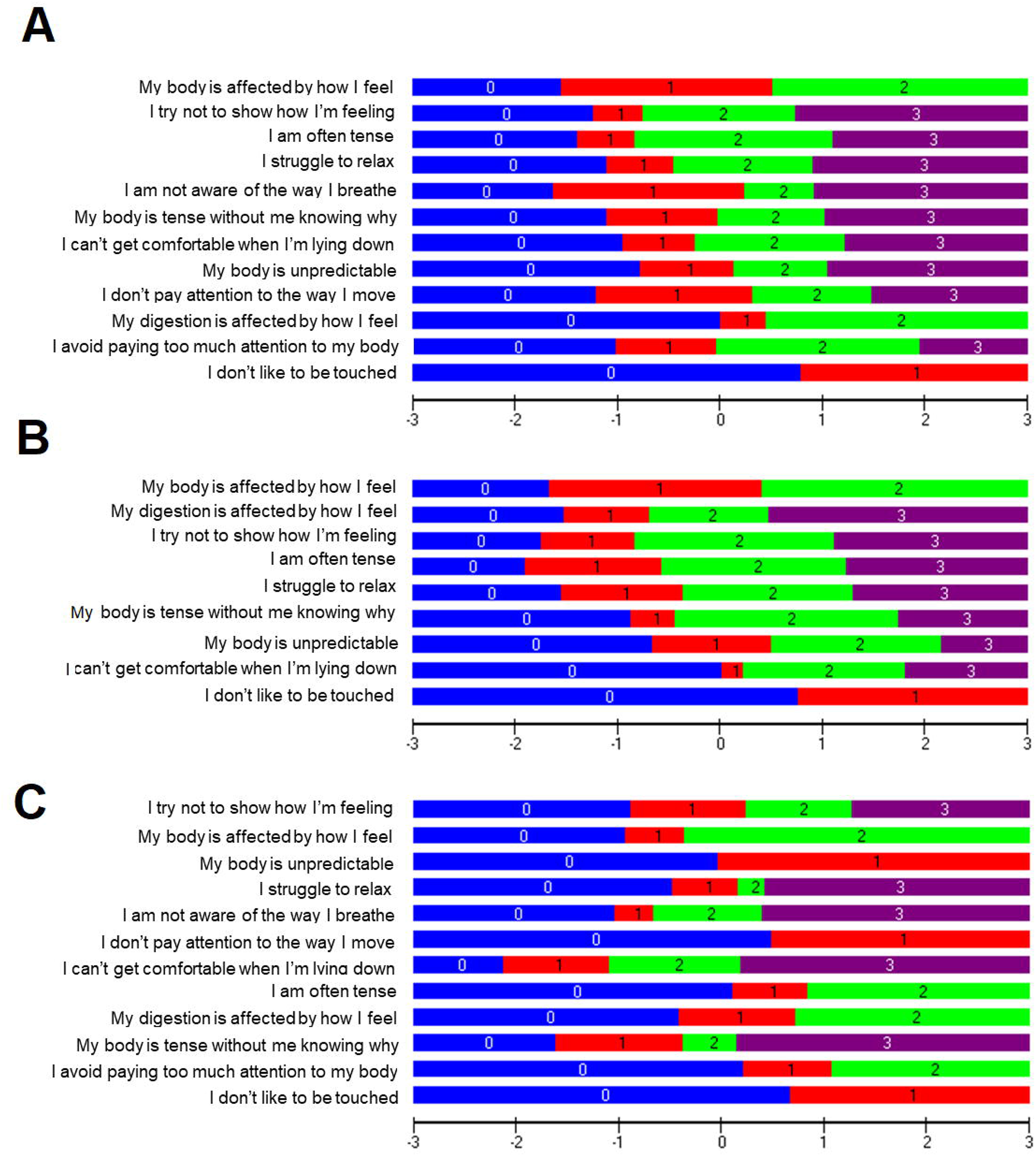
Item threshold map. ***Legend:*** The item threshold map shows the easiest items on the top to the hardest items on the bottom for **A:** Adults with musculoskeletal pain, **B:** community-dwelling adults, and **C:** adults with SCI and neuropathic pain. Using an interval logit scale, the horizontal black line at the bottom represents the location of the thresholds between the scoring as well as the body awareness ability level (higher scores indicating lower body awareness ability) of each participant.

### Community-dwelling adults

Initially, for community-dwelling adults, Rasch analysis of the BARQ-R showed that 1 item (item 11) presented misfit (Res=2.77, *p*=.000004) and 2 items (items 2,12) had disordered thresholds. Upon rescoring item 2 (0012) and item 12 (0011), there were no more disordered thresholds, but item 11 was still misfitting. After deleting item 11 (Res=3.03, *p*=.000004), item 3 (Res=3.33, *p*=.000092) and item 4 (Res=2.89, *p*=.00016) displayed misfit. After deleting the 2 items (items 3,4), the scale had no more items with disordered thresholds or misfitting items. There were only 4 (0.85%) misfitting persons. The person mean location was -0.62±1.09 logits (**Fig. 1B**) which is close to good targeting. There were minimal floor (2.63%), and ceiling effects (0.425%). The scale showed sufficient reliability for group decision-making in this population (PSR=0.74). No consequential LID was found. PCAR had an eigenvalue of 1.88 (percent variance 20.85%) but the paired *t*-test resulted in only 3.61% of persons having a significantly different logit location on the two subtests. This result is indicative of the scale being unidimensional. Similar to the group with musculoskeletal pain, “*My body is affected by how I feel*” was the easiest item for adults in the community, and “*I don’t like to be touched*” was the hardest item for this group (**Fig. 2B**). The score-to-measure table is provided in **Table 5**.

**Table 5.** Rasch-based score-to-measure tables for community-dwelling adults.

| Total<br>BARQ-R<br>Score | Logit | Converted<br>logits to 0-100 |
| --- | --- | --- |
| 0 | -3.87 | 0.00 |
| 1 | -3.05 | 10.40 |
| 2 | -2.48 | 17.66 |
| 3 | -2.08 | 22.71 |
| 4 | -1.77 | 26.71 |
| 5 | -1.50 | 30.11 |
| 6 | -1.27 | 33.13 |
| 7 | -1.05 | 35.90 |
| 8 | -0.85 | 38.50 |
| 9 | -0.65 | 40.99 |
| 10 | -0.46 | 43.41 |
| 11 | -0.27 | 45.79 |
| 12 | -0.09 | 48.18 |
| 13 | 0.10 | 50.57 |
| 14 | 0.29 | 53.02 |
| 15 | 0.49 | 55.55 |
| 16 | 0.70 | 58.21 |
| 17 | 0.92 | 61.03 |
| 18 | 1.16 | 64.09 |
| 19 | 1.43 | 67.45 |
| 20 | 1.73 | 71.26 |
| 21 | 2.08 | 75.74 |
| 22 | 2.51 | 81.31 |
| 23 | 3.12 | 89.10 |
| 24 | 3.98 | 100.00 |

### Adults with SCI-related neuropathic pain

Initially, for adults with SCI-related neuropathic pain, one item (item 12) showed misfit (Res=2.56, *p*=.00012) and 5 items (items 6, 8, 9, 11, and 12) had disordered thresholds. Upon rescoring items 6 (0011), 8 (0012), 9 (0012), 11 (0012), and 12 (0011), there were no misfitting items, however, there were 2 items (items 2, 3) with disordered thresholds. After rescoring item 2 (0012) and item 3 (0011), the scale had no more disordered thresholds and no misfitting items. There were no misfitting persons. The scale showed good targeting for adults with SCI and neuropathic pain (person mean location: -0.33±0.71 logits, **Fig. 1C**), no floor effect (0.00%), or ceiling effect (0.00%). However, the scale did not show sufficient reliability for this group (PSR=0.65). LID was found in 10 item pairs (**Table 3**). PCAR had an eigenvalue of 2.48 (20.66%), however, the paired t-test revealed only 4.55% of persons having a significantly different location on the two subtests, which points to the unidimensionality of the scale for this group. “*I try not to show how I’m feeling*” was the easiest item and “*I don’t like to be touched*” was the hardest item (**Fig. 2C**).

## DISCUSSION

For adults with self-reported musculoskeletal pain, we validated Dragesund *et al*. (2018)’s findings in a larger sample. The BARQ-R showed good item fit, good targeting, and reliability, which indicates that this scale is considered unidimensional and can be used for group decision-making, given that the PSR is above .70.

For community-dwelling adults with no reported pain, the targeting is close to the 0.5 logit limit, indicating that the items were only slightly too easy for this group. This result is not surprising given that this group does not have any known body awareness deficits and thus could provide a baseline level of what the range of body awareness ability is in a general healthy US community. The BARQ-R showed good item fit and reliability with PSR above .70. Therefore, this scale can be recommended for use in community-dwelling adults.

The preliminary Rasch analysis in adults with SCI-related neuropathic pain demonstrated that the BARQ-R had good item fit and good targeting. However, the reliability was too low, given that the PSR was below .70. Our results should also be validated in a larger sample before any recommendations can be made for use of this scale in adults with SCI.

Regarding unidimensionality, the paired *t*-tests indicated that the scale was unidimensional for community-dwelling adults and adults with SCI-related neuropathic pain. However, there may be multiple underlying aspects of body awareness in the BARQ-R for adults with self-reported musculoskeletal pain.

As noted by the PCAR, the following items showed positive loadings on the first component: “*I avoid paying too much attention to my body*,” “*I am not aware of the way I breathe*,” and “*I don’t pay attention to the way I move*.” The following items showed negative loadings on the first component: “I am often tense,” “*My body is affected by how I feel*,” “*I struggle to relax*,” and “*My body is tense without me knowing why*.” Conceptually, these two groupings may indicate two underlying aspects of body awareness. For example, all items with positive loadings seem to have negatively worded items and indicate a lack of awareness. This may be consistent with the “awareness” category in the original BARQ (Dragesund *et al*. 2010). Whereas all items with negative loadings on the first component may be measuring an aspect of being tense.

Across all groups, the hierarchy of items showed similar ‘easy’ items. Both adults with musculoskeletal pain and community-dwelling adults had the item “*my body is affected by how I feel”* as the easiest. Adults with SCI-related neuropathic pain had this item as the second easiest. Dragesund *et al*. (2018) also reported this item as the second easiest one. All groups had the same hardest item “*I don’t like to be touched*”, which aligns with Dragesund *et al*. (2018)’s findings as well.

Given the importance of body awareness training in clinical settings, it is critical to further examine the conditional minimally detectable change (cMDC) for the BARQ-R. In contrast to the MDC, which is a single index, the cMDC is a matrix of values for each pair-wise combination of change scores.^25^

### Limitations

Future studies should validate our preliminary findings in adults with SCI-related neuropathic pain. One limitation across all samples may be a lack of diversity in our samples. Our data was collected in adults in Minnesota, USA, where there is an 82.85% White population and 6.41% Black or African American.^26^ Therefore, future studies should attempt at recruiting a more racially and geographically diverse sample.

## CONCLUSIONS

Overall, the BARQ-R showed good item and person fit and proved to be sufficient for group decision-making in a clinical setting for both adults with musculoskeletal pain and community-dwelling adults. However, due to the low reliability and the fact that we only have preliminary results of the BARQ-R in adults with SCI-related neuropathic pain, we cannot yet recommend the BARQ-R for clinical use in adults with SCI until a further Rash analysis is carried out in a larger sample and the reliability is higher.

Future research should also focus on validating these findings in a more inclusive population as well as in other populations that may have body awareness deficits.

## Data Availability

All data produced in the present study are available upon reasonable request to the authors.

## Abbreviation List

BARQ: Body Awareness Rating Questionnaire
BARQ-R: Revised Body Awareness Rating Questionnaire
cMDC: conditional Minimally Detectable Change
NPMP: Norwegian Psychomotor Physiotherapy
LID: Local item dependence
PSR: Personal Separation Reliability
RULER: Rasch Reporting Guideline for Rehabilitation Research
RUMM2030: Rasch Unidimensional Measurement Model
SCI: Spinal Cord Injury
US: United States

## Acknowledgments

We thank all participants; the Driven to Discover (D2D) Research Team; and the volunteers who helped at the D2D Facility at the Minnesota State Fair and at Highland Fest for providing us with the opportunity to carry out this study. We would like to express our profound thanks to Marc Noël for the critical review of the manuscript.

## Author Contributions

AVDW designed the project. AVDW, WD, and other volunteers enrolled participants and collected the data. AVDW analyzed and interpreted the data sets. SC wrote the manuscript. AVDW, WD, and JB provided crucial revisions and edits to the manuscript. Every author takes responsibility for the final draft.

---

STROBE Statement—Checklist of items that should be included in reports of ***cross-sectional studies***

|  | Item No | Recommendation | Page No |
| --- | --- | --- | --- |
| Title and abstract | 1 | (a) Indicate the study’s design with a commonly used term in the title or the abstract | 1 |
|  |  | (b) Provide in the abstract an informative and balanced summary of what was done and what was found | 2-3 |
| Introduction |  |  |  |
| Background/rationale | 2 | Explain the scientific background and rationale for the investigation being reported | 4-5 |
| Objectives | 3 | State specific objectives, including any prespecified hypotheses | 5 |
| Methods |  |  |  |
| Study design | 4 | Present key elements of study design early in the paper | 5 |
| Setting | 5 | Describe the setting, locations, and relevant dates, including periods of recruitment, exposure, follow-up, and data collection | 5 |
| Participants | 6 | (a) Give the eligibility criteria, and the sources and methods of selection of participants | 5-6 |
| Variables | 7 | Clearly define all outcomes, exposures, predictors, potential confounders, and effect modifiers. Give diagnostic criteria, if applicable | 6-7 |
| Data sources/ measurement | 8* | For each variable of interest, give sources of data and details of methods of assessment (measurement). Describe comparability of assessment methods if there is more than one group | 6-7 |
| Bias | 9 | Describe any efforts to address potential sources of bias | N/A |
| Study size | 10 | Explain how the study size was arrived at | N/A |
| Quantitative variables | 11 | Explain how quantitative variables were handled in the analyses. If applicable, describe which groupings were chosen and why | N/A |
| Statistical methods | 12 | (a) Describe all statistical methods, including those used to control for confounding | 6-7 |
|  |  | (b) Describe any methods used to examine subgroups and interactions | N/A |
|  |  | (c) Explain how missing data were addressed | N/A |
|  |  | (d) If applicable, describe analytical methods taking account of sampling strategy | N/A |
|  |  | (e) Describe any sensitivity analyses | N/A |
| Results |  |  |  |
| Participants | 13* | (a) Report numbers of individuals at each stage of study—eg numbers potentially eligible, examined for eligibility, confirmed eligible, included in the study, completing follow-up, and analysed | N/A |
|  |  | (b) Give reasons for non-participation at each stage | N/A |
|  |  | (c) Consider use of a flow diagram | N/A |
| Descriptive data | 14* | (a) Give characteristics of study participants (eg demographic, clinical, social) and information on exposures and potential confounders | 7 |
|  |  | (b) Indicate number of participants with missing data for each variable of interest | N/A |
| Outcome data | 15* | Report numbers of outcome events or summary measures | 8-9<br>Table 1<br>Table 2<br>Table 3 |
|  |  |  | Table 4 |
|  |  |  | Table 5 |
| Main results | 16 | (a) Give unadjusted estimates and, if applicable, confounder-adjusted estimates and their precision (eg, 95% confidence interval). Make clear which confounders were adjusted for and why they were included | N/A |
|  |  | (b) Report category boundaries when continuous variables were categorized | N/A |
|  |  | (c) If relevant, consider translating estimates of relative risk into absolute risk for a meaningful time period | N/A |
| Other analyses | 17 | Report other analyses done—eg analyses of subgroups and interactions, and sensitivity analyses | N/A |
| **Discussion** |  |  |  |
| Key results | 18 | Summarise key results with reference to study objectives | 9-11 |
| Limitations | 19 | Discuss limitations of the study, taking into account sources of potential bias or imprecision. Discuss both direction and magnitude of any potential bias | 11 |
| Interpretation | 20 | Give a cautious overall interpretation of results considering objectives, limitations, multiplicity of analyses, results from similar studies, and other relevant evidence | 11-12 |
| Generalisability | 21 | Discuss the generalisability (external validity) of the study results | 12 |
| **Other information** |  |  |  |
| Funding | 22 | Give the source of funding and the role of the funders for the present study and, if applicable, for the original study on which the present article is based | 12 |

\*Give information separately for exposed and unexposed groups.
**Note:** An Explanation and Elaboration article discusses each checklist item and gives methodological background and published examples of transparent reporting. The STROBE checklist is best used in conjunction with this article (freely available on the Web sites of PLoS Medicine at http://www.plosmedicine.org/, Annals of Internal Medicine at http://www.annals.org/, and Epidemiology at http://www.epidem.com/). Information on the STROBE Initiative is available at www.strobe-statement.org.

